# The unique face of anxious depression: Exaggerated threat but preserved positive valence reactivity

**DOI:** 10.1101/2022.05.12.22275025

**Authors:** Maria Ironside, Rayus Kuplicki, Ebony A. Walker, Tate Poplin, Cheldyn M. Ramsey, Katherine L. Forthman, Melissa E. Nestor, Robin L. Aupperle, Salvador M. Guinjoan, Sahib S. Khalsa, Jonathan Savitz, Jennifer L. Stewart, Teresa A. Victor, Martin P. Paulus

## Abstract

**Background:** Even though anxious depression is among the most prevalent psychiatric conditions; its underlying neural and behavioral characteristics remain not well understood. This may be important to break down heterogeneity in depression. This study investigated the unique profile of individuals with anxious depression using affective startle modulation, a process known to independently probe appetitive/defensive systems and known to be affected by mood and anxiety disorders

**Methods:** 236 depressed participants of the Tulsa 1000 study completed multi-level assessments including an emotional reactivity task with eye-blink startle measurement. To minimize bias due to covariates, 124 participants with comorbid depression and anxiety disorders (Dep+Anx) were matched with 62 participants with depression only (Dep). Eye-blink startle magnitudes during positive/negative visual cues were analyzed.

**Results:** The Dep group showed no affective modulation of startle. However, the Dep+Anx group showed potentiation from aversive cues and attenuation from appetitive cues. The Dep+Anx group also showed increased attenuation from appetitive cues compared to the Dep group. Dimensionally, the effect of self-report anxiety on startle was moderated by self-report depression.

**Conclusions:** Compared to individuals with depression, those with anxious depression demonstrate heightened positive/negative startle modulation, with depression levels moderating the link between anxiety sensitivity and startle reflex. The differences between these groups in processing aversive/appetitive information support the conclusion that these depression subtypes should be considered separately in future clinical trials. ClinicalTrials.gov identifier: #NCT02450240.

**General scientific summary:** This study suggests that there are striking differences between those with anxious and non-anxious depression. Specifically, these two groups differ in terms of threat reactivity measured by eye-blink startle response. This proposes that the two groups be separated in future clinical trials.

## Introduction

Nearly half of individuals with Major Depressive Disorder (MDD) also have an anxiety disorder (Kessler et al., 2015). Thus, comorbid MDD and anxiety disorders (e.g. generalized anxiety disorder, social phobia, panic disorder, or simple phobia) are among the most common presentations for mental health providers. Yet, the underlying brain and behavioral processes that characterize anxious depression are still incompletely understood. A better understanding of the underlying process dysfunctions is an important first step in the development of more mechanistically oriented interventions. Anxious depression is associated with greater treatment resistance, (Ionescu et al., 2014), quicker symptom relapse (Fava et al., 2008), poorer treatment outcomes, (Penninx et al., 2011) and higher levels of suicidal ideation (Kessler et al., 2015) than non-anxious depression. However, treatment engagement is reported to be higher (Kessler et al., 2015). Typical treatments include traditional antidepressants augmented with benzodiazepines or atypical antipsychotics. This can be problematic because of the risk for use disorder and unfavorable side effects profiles, which can be increased in anxious depression (Gaspersz et al., 2017).

MDD and anxiety have some overlapping dysfunction in neurocognitive processes (McTeague et al., 2020). However, there are important features which make them distinct. According to Clark and Watson’s seminal model of depression and anxiety hyperarousal is a key feature of anxiety and blunted positive responding or anhedonia is a key feature of depression (Clark & Watson, 1991). Adding to this, the emotion context insensitivity model (Rottenberg et al., 2005) reports attenuation of both positive and negative affect in MDD, proposing a general level of emotional blunting in MDD. It is unclear what happens when this emotional blunting and hyperarousal are combined in anxious MDD. Additionally, there is presumably a healthy level of “arousal” that lies between blunted non-anxious MDD and “hyperaroused” anxiety/anxious MDD. This means that when those with anxious and non-anxious MDD are merged into a single clinical sample, comparisons on measures of arousal with healthy people are not helpful, as these two clinical groups lie on either end of the arousal spectrum. Furthermore, studies comparing anxious versus non-anxious MDD are rare, which may be impeding treatment selection and development.

Following on from this, hyperarousal is a feature of anxiety and anxious MDD, that is not likely to be present in non-anxious depression and therefore may be a useful candidate target mechanism. Hyperarousal in anxious MDD is associated with dysregulation of stress circuitry (Powers et al., 2016) and cortical thinning in prefrontal areas associated with top-down aspects of emotional regulation (Zhao et al., 2017). Threat responses are particularly associated with anxiety (Bar-Haim et al., 2007) and have been shown to engage a corticolimbic circuit (Mobbs et al., 2009; Mobbs et al., 2007). In a functional magnetic resonance imaging (fMRI) study comparing anxious and non-anxious MDD, limbic responses to emotional conflict (presentation of emotional facial expressions and non-task relevant incongruent emotional words) were similar across anxiety and MDD but prefrontal regulation of emotional conflict was absent in anxiety/ anxious MDD but preserved in non-anxious MDD (Etkin & Schatzberg, 2011), suggesting that deficits in top-down aspects of emotional regulation may be key to this comorbidity. Emotional regulation is associated with structural and functional connectivity of frontal and limbic regions (Kim et al., 2011). MDD is associated with reduced corticolimbic functional connectivity (Kaiser et al., 2015), although findings are mixed (Williams, 2017), perhaps due to the fact that MDD is comprised of heterogeneous subgroups. Examining the anxious MDD subgroup may remove some of the variance, but few studies have examined this. In a study of depressed patients, amygdala functional connectivity mediated the relationship between anxiety and MDD (He et al., 2019). Together, this suggests that top-down control of defensive responses to acute threat (and the associated circuits) may be a suitable cognitive target specific to anxious versus non-anxious MDD.

The startle reflex is a defensive eyeblink response to an intense stimulus. The plasticity of this reflex in relation to positive or negative affective context makes this an ideal experimental approach to examine alterations of the defense system (Boecker & Pauli, 2019). In healthy participants startle reflex is attenuated by appetitive stimuli and potentiated by aversive stimuli; a phenomenon called affective startle modulation (ASM) (Bradley et al., 1990; Lang et al., 1990); explained by motivational priming of the appetitive/defensive systems (Lang, 1995). Hereby congruent aversive motivational states prime the defensive system, potentiating the defensive startle response, whereas incongruent appetitive states attenuate the response. Deviations from the expected pattern of ASM are thought to reflect abnormal functioning of the underlying motivational system. A recent review of ASM and psychopathology (Boecker & Pauli, 2019) suggests that the defensive/appetitive systems operate independently and characterize depressed and anxious psychopathology in the following ways: 1) *increased* affective startle potentiation (ASP) to aversive stimuli in anxiety (Cook et al., 1992; Cook et al., 1991; Garner et al., 2011; Temple & Cook, 2007) and to phobic stimuli in phobia (Globisch et al., 1999), and 2) *general hyporeactivity* to aversive and appetitive affective stimuli in MDD, with blunted affective startle modulation in severe MDD (Kaviani et al., 2004). Interestingly, in a study examining a range of anxiety patients, responses were blunted in participants with more pervasive disorders such as multiple trauma PTSD and co-morbidity with MDD (McTeague & Lang, 2012), suggesting disengagement of the defense system in response to chronic disease course. These opposing forces of depression and anxiety on startle make the examination of anxious MDD necessary and potentially useful. In practical terms, startle paradigms are more clinically feasible to implement than neuroimaging and have recently been employed as a way to screen novel anxiolytics (Grillon & Ernst, 2020).

Preclinical literature has established a key role for the defensive and appetitive neuronal systems in the modulation of startle reflex, namely the bed nucleus of the stria terminalis (BNST), the amygdala (Davis et al., 1999) and the nucleus accumbens (Koch et al., 1996). Concurrent neuroimaging and startle electrophysiology is technically difficult, historically limiting human findings to lesion studies of the amygdala showing elimination of negative ASM (Buchanan et al., 2004). However, recent advances allowed converging evidence in human fMRI of valence-specific triggered amygdala responding, with individual level associations between startle magnitude and neural activation strength, suggesting that startle measures could be used as a direct read-out of neural activation of the central amygdala (Kuhn et al., 2020), allowing measurement of a key component of the corticolimbic circuit implicated in anxious MDD.

ASM to aversive stimuli is dimensionally associated with trait fear (Vaidyanathan et al., 2009), a self-report measure of threat sensitivity. The goal of this investigation was to determine whether anxious MDD shows a unique profile of exaggerated defense related processes (i.e. hyperarousal) using data from a large transdiagnostic sample collected as part of the Tulsa 1000 study (Victor et al., 2018). The basic approach was to compare a propensity-matched sample of depressed and anxious depressed individuals using a multi-level approach focused on positive/ negative valence and threat using symptoms and physiological levels of analysis. Based on previous findings, we hypothesized that individuals with anxious depression but not those with non-anxious depression would show an increased threat-related startle response pattern characterized by an altered ASM response to negative stimuli compared to positive stimuli.

## Methods and Materials

### Participants

Data were collected from 236 participants (170 female) from the Tulsa 1000 study (Victor et al., 2018), a naturalistic longitudinal study recruiting a community. Participants were between 18 and 56 years of age at the time of electromyography (EMG) measurements (mean age = 35.6, standard deviation = 11.4). Participants were screened for inclusion on the basis of the Patient Health Questionnaire (PHQ-9) ≥10. Exclusion criteria were positive urine drug screen; lifetime bipolar, schizophrenia spectrum, antisocial personality, or obsessive compulsive disorders; active suicidal ideation with intent or plan; moderate to severe traumatic brain injury; severe and or unstable medical concerns; changes in psychiatric medication dose in the last 6 weeks; and fMRI contraindications. Ethical approval was obtained from Western Institutional Review Board T1000 protocol #20142082. Full exclusion criteria can be found in the supplement and the parent project protocol paper (Victor et al., 2018). All participants provided written informed consent prior to participation, in accordance with the Declaration of Helsinki, and were compensated for participation. ClinicalTrials.gov identifier: #NCT02450240.

After removal of 24 participants with greater than 20% unusable EMG data (see supplement for CONSORT diagram and below for exclusion criteria) and one participant for incomplete self-report data the initial sample for the analysis included 62 participants with non-anxious MDD (Dep) and 149 participants with comorbid MDD and anxiety disorder (Dep+Anx), defined categorically as lifetime MDD and at least one anxiety disorder according to the anxiety module of the Mini International Neuropsychiatric Interview (MINI; (Sheehan et al., 1998)), these include Generalized Anxiety Disorder, Panic Disorder, Agoraphobia and Social Phobia (see Table 1 for details). To reduce the bias due to confounding variables, 124 participants from the Dep+Anx group were propensity matched for age, sex and education at a ratio of 2:1 with 62 participants from the Dep group using the MatchIt package in R (R Core Team, 2020). Propensity score analysis is based on the hypothesis that two patients with similar propensity scores have covariates which come from similar distributions. This means that by selecting or reweighting samples based on propensity scores, researchers create new datasets where covariates are similar between two groups (Zhao et al., 2021). These two groups did not differ on their level of depression (PROMIS) (see **Table 1**) but, crucially, had significantly different levels of trait self-report anxiety sensitivity (ASI) and state anxiety severity and impairment (OASIS).

**Table 1:**
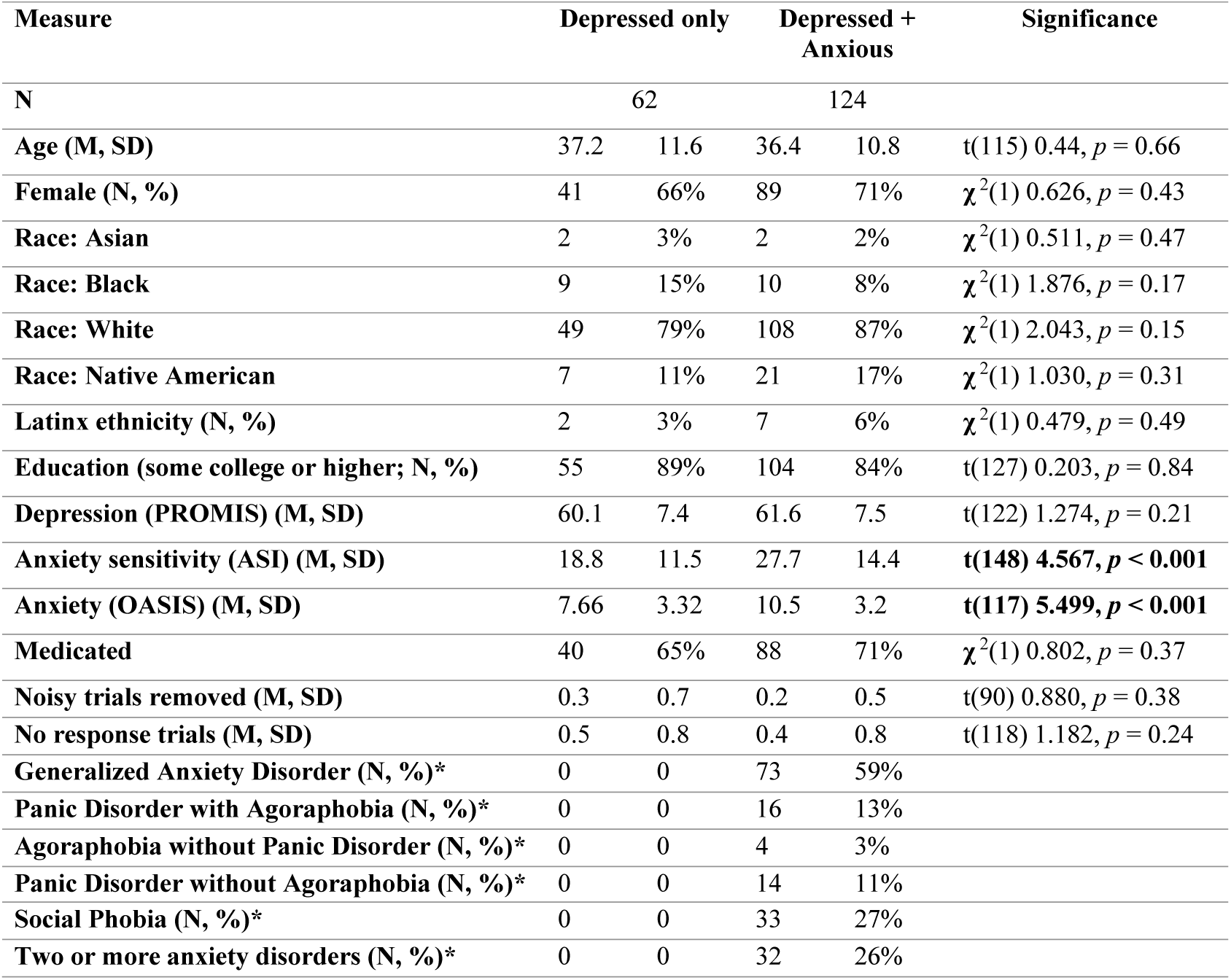
Final sample demographics. M: Mean; SD; standard deviation; PROMIS: Patient-Reported Outcomes Measurement Information System; ASI: Anxiety Sensitivity Index; OASIS: Overall Anxiety Symptom and Impairment Scale * Anxiety disorders as defined by the clinician administered Mini International Neuropsychiatric Interview (MINI; (Sheehan et al., 1998))

### Procedure

General procedures included a clinical interview session, a neuroimaging session and a behavioral (electrophysiology) session, completed within two weeks on average. Although the parent project (i.e., T1000) consisted of a broader range of protocols, only details relevant to the current study are presented here. See protocol paper (Victor et al., 2018) for full details.

Study staff administered the MINI clinical interview. During this session, participants also provided self-reported information on demographics. For the current study we focused on dimensional measures of anxiety/threat sensitivity (Anxiety Sensitivity Index; ASI), depression (PROMIS depression scale) and as a follow up, approach and avoidance motivation (Behavioral Inhibition/ Behavioral Activation scale; BIS/BAS).

Participants completed an emotional reactivity task (Lang et al., 1990) during EMG eyeblink recording (**Fig. 1**). Facial EMG was recorded to assess physiological startle from the orbicularis oculi in accordance with published guidelines (Blumenthal et al., 2005). Two electrodes were attached below the lower eyelid of the left eye, one below the outer edge and one below the centre of the eye (distance approx. 15 mm). Participants viewed appetitive, neutral, and aversive images from the International Affective picture series (IAPS) (Lang et al., 1999) for 6 s. Noise probes (95 dB) were presented between 2500 and 4500 ms after picture onset to elicit startle blink responses during 24 of these image presentations (8 per valence). Electrocardiogram (EKG) data were also collected at a baseline session (during rest) and during the emotional reactivity task. Analyses were carried out on the first 500 participants of the T1000 sample and were not preregistered. Confirmatory analyses from the second 500 participants will be preregistered.

**Figure 1:**
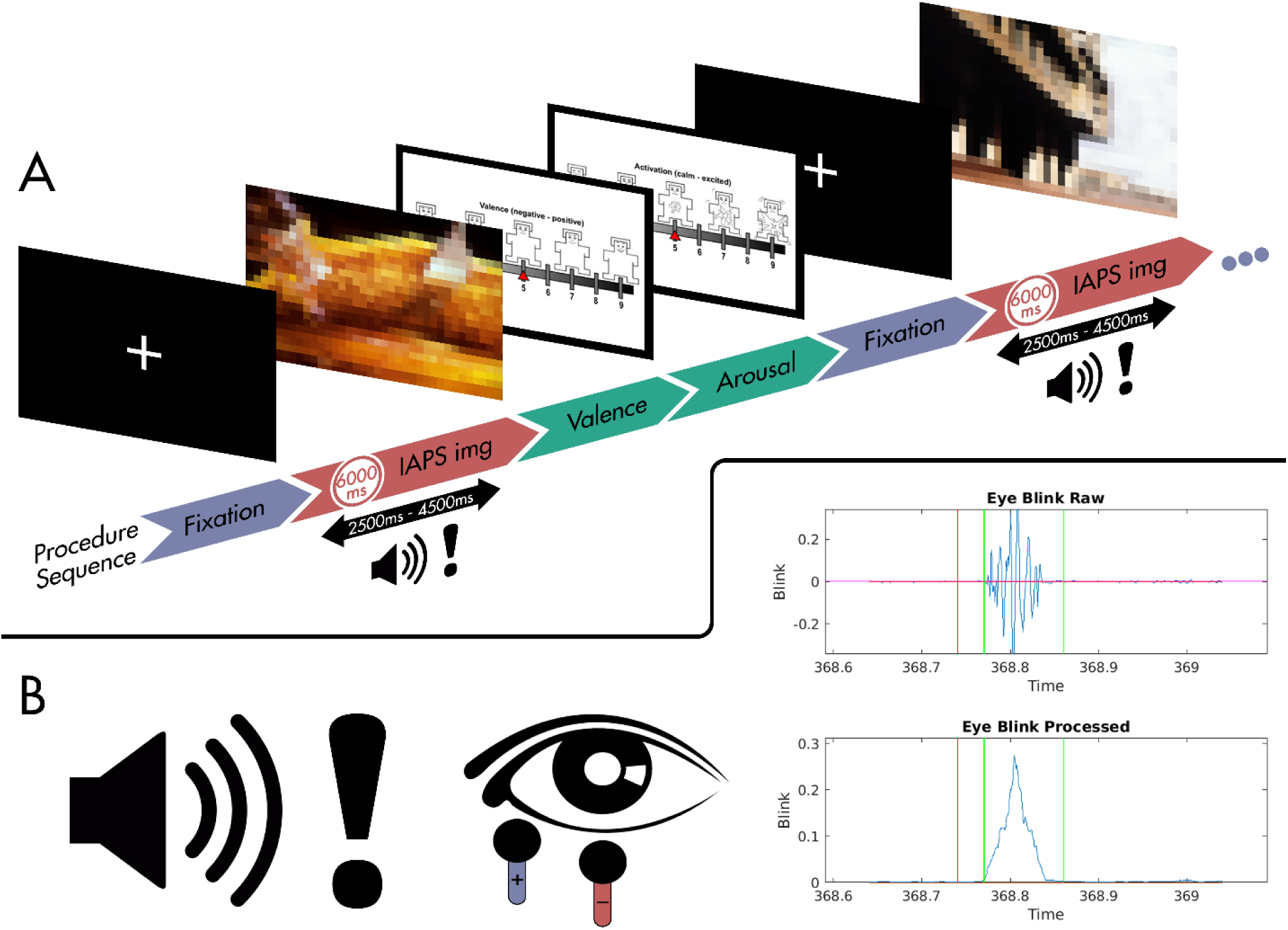
Emotional reactivity task: A) Each trial begins with a 20-26s fixation period, followed by presentation of one image for 6s, during which a startle probe is presented. After each image, the participant makes valence and arousal ratings on a 7 point scale. (IAPS images blurred in this schematic for copyright reasons; first example image is a pancake (appetitive), second example image is a building (neutral)) B) Startle EMG data collected from electrodes placed on the orbicularis oculi.

### Data pre-processing and analysis

#### EMG data

EMG data were analyzed following accepted guidelines (Blumenthal et al., 2005). Briefly, raw data were bandpass Butterworth filtered between 60 and 500 Hz, smoothed over every rolling 20 samples with a resolution of 0.0005 sec. Quality control of the data was carried out using automated processes implemented in Matlab R2019a (MATLAB) and visual inspection. For the automated process “bad” trials were excluded where the standard deviation of the baseline (100 msec before noise stimuli) was greater than 2x the standard deviation of the response window during the trial (30-90 msec post noise stimuli). Any blinks occurring after the noise stimuli but before the beginning of the response window were excluded. Startle magnitude was calculated as the difference between the maximum magnitude in the response window minus the mean baseline. No blink trials were those in which the peak value in the response window is smaller than the range of the baseline and were included in the startle averages (Blumenthal et al., 2005). Individual trials and videos of electrode placement were visually inspected by two investigators (EW and CR) and additional trials/participants were excluded if electrode positioning was poor or if there were artifacts in the signal. Data were positively skewed. Therefore, for analyses examining valence interactions t-scores were calculated for each trial using a within-participant formula:

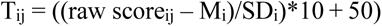

For analyses using raw startle reflex only an optimized log transform (Forthman, 2019) was used to counteract skew. For repeated measures, data were analyzed with mixed effects linear regression using the lmer package in R (R Core Team, 2020) with fixed factors of valence and group, a within participant adjustment for the slope of valence (random slope) and covariates of sex and age. Degrees of freedom were estimated using Satterthwaite’s method. For non-repeated measures data were analyzed with linear regression using the stats package in R. Dimensional analyses examined the interaction effect of anxiety sensitivity and depression on startle.

#### EKG data

EKG data were partitioned into 5 minute blocks following accepted guidelines (Shaffer & Ginsberg, 2017). Heart rate variability metrics were calculated using HRVTool (Vollmer, 2019) in Matlab (MATLAB). High frequency and low frequency band power were extracted as frequency domain measures for parasympathetic (rest and digest) and sympathetic (fight or flight) activity. In addition root mean square of successive differences (RMSSD) and standard deviation of the N-N intervals (SDNN) were calculated as time domain measures of overall heart rate variability (HRV). Effects of group and task were analyzed with mixed effects linear regression using the lmer package in R (R Core Team, 2020) with fixed factors of task (versus baseline) and group, a within participant adjustment (random intercept) and covariates of sex and age.

## Results

### Startle response

Mixed effects linear regression results showed a significant *valence X group* interaction (F(2,310) 3.460, *p* = 0.03, *R*^2^ = 0.022). Planned contrasts showed that the Dep group had no modulation of startle response from positive or negative valence (pairwise comparisons; all *p* > 0.5), whereas the Dep+Anx group showed negative potentiation compared to positive (negative > positive) (t(184) = 4.608, *p* < 0.001) and positive attenuation compared to neutral (positive < neutral) (t(548) = -5.018, *p* < 0.001) (**Fig. 2**). The Dep+Anx group also had lower startle response during appetitive stimuli compared to the Dep group (t(397) = -2.273, *p* = 0.02). Supplemental analyses comparing to healthy controls are included for completeness in the Supplement.

**Figure 2:**
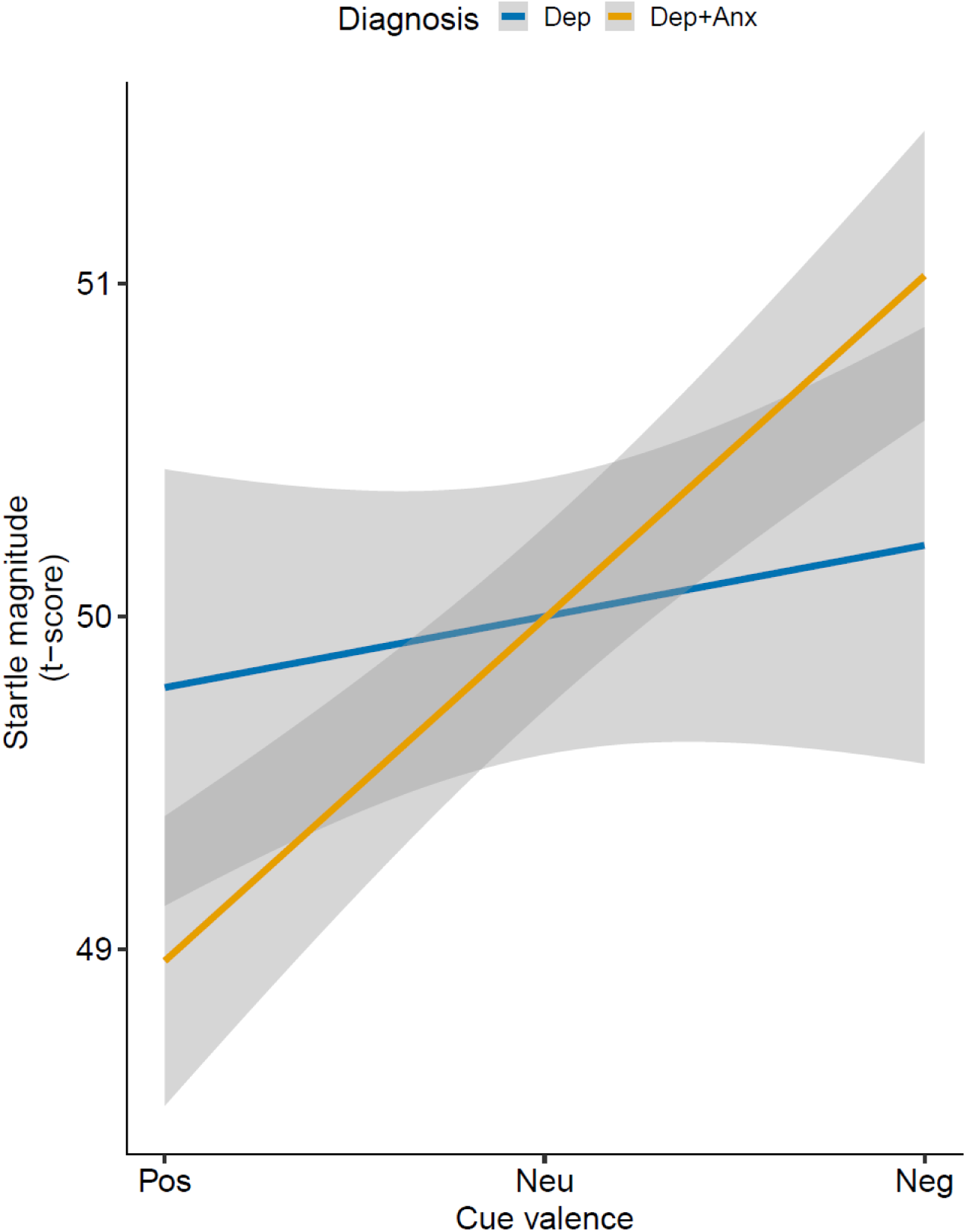
Blunted affective startle modulation in depression versus comorbid depression and anxiety. Shaded area represents the 95% confidence interval.

### Image ratings

Participants rated the aversive images as more negatively valenced (t(368) = 37.948, *p* < 0.0001) and more arousing (t(368) = 16.898, *p* < 0.0001); and the appetitive images as more positively valenced (t(368) = 15.033, *p* < 0.0001) and more arousing (t(371) = 3.354, *p* < 0.001) than the neutral images. There was no *valence* X *group* interaction on ratings (F(2,368) 0.173, *p* = 0.84). RTs of arousal ratings had a significant *valence* X *group* interaction (F(2,368) 4.11, *p* = 0.03), driven by slower RTs of arousal ratings for aversive images in the Dep group compared to the Dep+Anx group (t(295) = 2.902, *p* = 0.004) (Supplemental Fig. S3). There were no main effects of group on RTs for valence or arousal ratings (all *p* > 0.16).

### Dimensional analyses

Dimensional analyses in the same sample examined the effects of self-report anxiety sensitivity (ASI) and depression (PROMIS) on raw startle response using linear regression. There was a significant *ASI* X *Depression* interaction. This indicates that depression moderated the effect of anxiety sensitivity (F(1,178 = 4.609 *p* = 0.03, R^2^ = 0.032). For the lower three quartiles of self-report depression, ASI was significantly correlated with overall startle reflex (r(112) = 0.23, *p* = 0.01), but this relationship was not observed in the top quartile (r(72) = -0.16, *p* = 0.18). This interaction was present when examining mean startle across all trials (**Fig. 3**) and during appetitive (*p* = 0.05) or aversive (*p* = 0.01) stimuli separately but not when examining difference scores of aversive versus appetitive or neutral (all *p* > 0.7).

**Figure 3:**
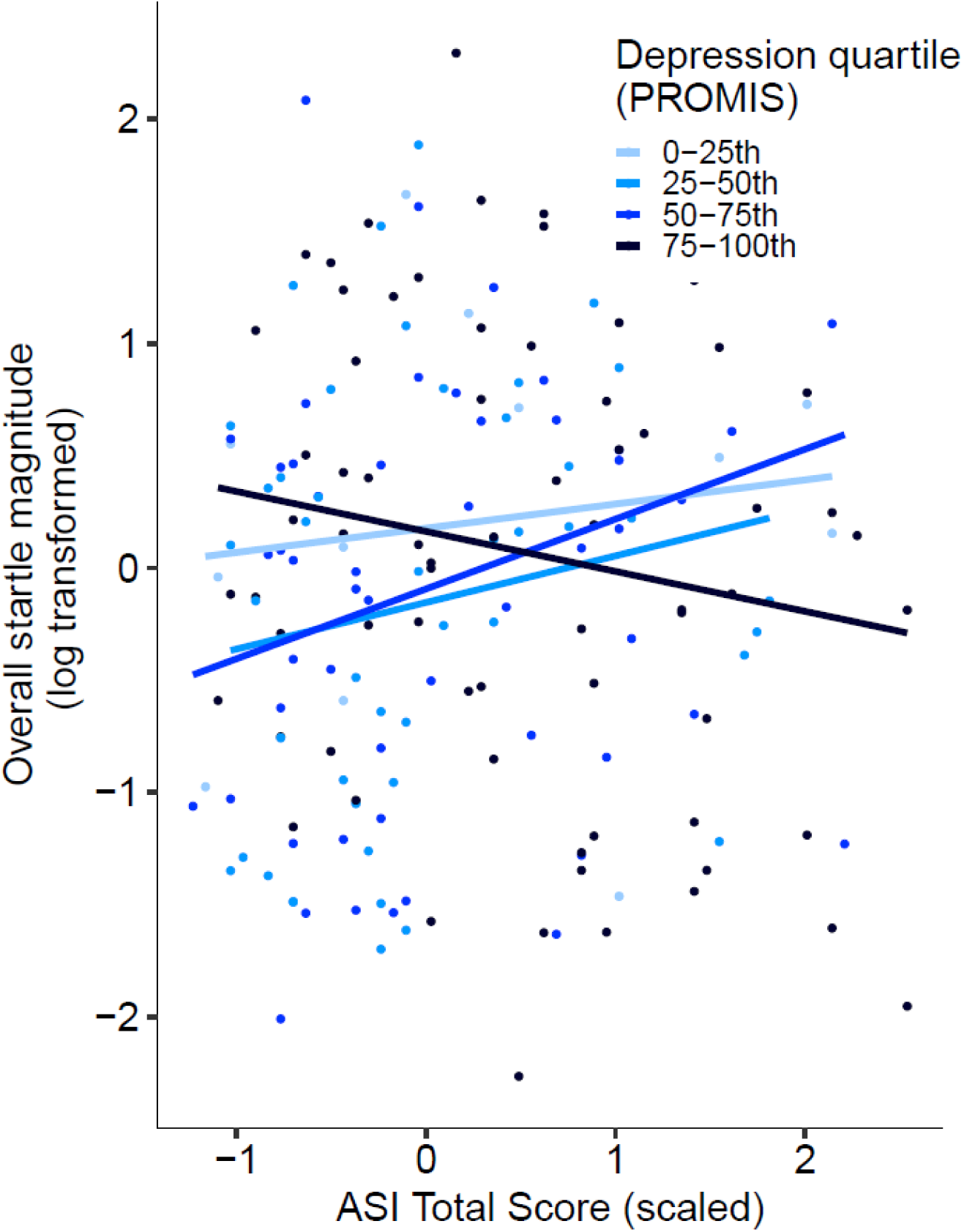
Moderation effect of self-report depression scores on effect of self-report anxiety sensitivity on overall startle response. PROMIS: Patient-Reported Outcomes Measurement Information System; ASI: Anxiety Sensitivity Index.

### Heart-rate variables

Low frequency HR power (F(1, 131) = 28.33, corrected *p* < 0.001) and SDNN (F(1, 131) = 11.44, corrected *p* = 0.004) were significantly increased during the emotional reactivity task compared to baseline resting state. There was no significant effect of group on low frequency HR power and SDNN or any effect of group or task on high frequency HR or RMSSD (all corrected *p* > 0.13).

### Follow-up analyses of behavioral approach and inhibition

To further characterize the groups they were compared on behavioral inhibition and approach using the BIS/BAS. A Welch two-sample t-test showed that the Dep+Anx group had higher behavioral inhibition (avoidance) than the Dep group (t(108) = 3.356, *p* < 0.001). There were no differences between the groups on behavioral activation (all *p* > 0.09).

## Discussion

This multi-level investigation aimed to examine whether anxious MDD is characterized by a unique profile of exaggerated defense related processes relative to propensity matched participants with non-anxious MDD. There were four main results: First, at a physiological level of analysis the Dep+Anx group showed preserved ASM whereas the Dep group showed no ASM; Second, at a behavioral level of analysis, the Dep group had slower RTs to arousal ratings for aversive cues than Dep+Anx; Finally, at a self-report level of analysis, depression moderated the effects of anxiety sensitivity on overall startle reflex where the most severely depressed individuals did not have an association between anxiety sensitivity and startle reflex. We further characterized the Dep+Anx group as having greater self-report behavioral avoidance. Together, these results support the hypothesis that anxious depression is characterized by affective modulation of appetitive and defensive systems relative to those with non-anxious depression and that non-anxious depression only is associated with valence independent blunting of emotional reactivity.

Previous work based on symptom patterns supports a tripartite model (Clark & Watson, 1991), consisting of a general negative affect factor and specific MDD and anxiety factors. The general factor is increased negative affect, common to both MDD and anxiety. In our findings this would be represented by the self-report depression scale which was similar in Dep versus Dep+Anx, showing that this general factor is associated with blunted overall startle. The specific MDD factor in the tripartite model is the absence of positive affect (anhedonia). In the emotional reactivity task the positively valenced images prime the appetitive system. As expected from the tripartite model, we see a lack of positive affective modulation from appetitive images in Dep. However, this was not the case in the Dep+Anx group, where we found preserved positive affective modulation. This suggests a novel key difference that may help explain greater treatment engagement in anxious depression (Kessler et al., 2015). The anxiety specific factor in the tripartite model is hyperarousal. In the emotional reactivity task the negatively valenced images prime the defensive system. This is illustrated by EKG analyses showing increased sympathetic activation in this task compared to rest. As expected from the tripartite model the Dep+Anx group show hyperarousal, represented by higher affective modulation from negative compared to positive images, which is not observed in Dep, suggesting another key difference. Hyperarousal is also represented by self-report anxiety sensitivity, which was also significantly higher in Dep+Anx versus Dep. These two measures of hyperarousal were significantly correlated in all but the most depressed participants, suggesting a moderating effect of depression.

This blunting of negative emotional reactivity aligns with the ECI model of MDD (Rottenberg et al., 2005), which seems not to apply to MDD when it is comorbid with anxiety. The striking difference between these two groups argues that they should not be combined for trials that target measures of hyperarousal, as differences from HCs will be obscured by the depressed and anxious clinical groups residing on either end of the “hyperarousal spectrum”. To illustrate this, supplemental analyses from a smaller HC group suggest that the HC ASM profile is between the Dep+Anx group and the Dep group. This may reflect a “normal” level of arousal between blunted Dep and hyperaroused Dep+Anx. This could be interpreted as anxiety “rescuing” the impairment from depression. However, this interpretation contrasts with the clinical observations that comorbid MDD and anxiety disorders are associated with worse clinical outcomes than MDD alone. Therefore, additional studies will be necessary to elaborate on the connection between startle dysfunction and clinical outcomes in individuals with Dep+Anx.

The approach-withdrawal model helps tease apart the behavioral factors, with depression being associated with reduced approach and anxiety being associated with increased avoidance (Davidson, 1998). In the present study, reduced approach and withdrawal could be compared to a lack of appetitive/approach or defensive/withdrawal priming in Dep, which is preserved in Dep+Anx. Speculatively, this suggests that treatments targeting the appetitive system (e.g. antidepressants acting on the dopamine system such as bupropion) may not be as effective in anxious depression, as (in terms of startle modulation at least) this system seems to be somewhat preserved. Conversely, treatments that target hyperarousal (e.g. GABAergic treatments) may not be necessary in non-anxious depression and may, we speculate, contribute towards further emotional blunting. Future research should examine how those with anxious versus non-anxious MDD respond to treatments separately targeting the appetitive and defensive systems.

These results add to prior work (Ironside et al., 2023) showing increases in interoceptive and nociceptive reactivity in anxious versus non-anxious MDD. Together, these findings have led us to propose a process model for anxious MDD to help distinguish disease modifiable processes that could be targeted with treatment. Anxiety may be characterized by excessive defense system activation which may be highly taxing and result in exhausting affective processing capacities ultimately resulting in MDD. This is in line with data showing that two-thirds of individuals with lifetime comorbid anxiety disorders and MDD reported an earlier age-of-onset of their anxiety disorder than their MDD (Kessler et al., 2015). In comparison, individuals with non-anxious MDD may be characterized by a primary lack of reactivity to affective stimuli, which results in a lack of anxious responding even when threat occurs. Therefore, whereas the primary disease-modifying process for anxious MDD would be to attenuate threat-related processing, the primary disease-modifying process for non-anxious MDD would be to enhance valence-related processing in general. Reward responsivity is a well-studied process in MDD, but we propose a more general factor of blunted valence responding, first proposed by the ECI model of MDD (Rottenberg et al., 2005) and supported by these data. This would modify the tripartite model, transforming the anxiety specific factor of hyperarousal into a general factor “arousal” with depression and anxiety working in opposition. If this is the case, the ECI model is insufficient to explain anxious MDD and further characterization is needed.

This study had several limitations. First, case-control designs and cross-sectional studies cannot arbitrate between cause and effect. Thus, mechanistic explanations require future experimental testing. Second, although the paradigm has been used extensively in prior investigations, compared to other experimental paradigms this version had fewer trials. Third, the Dep+Anx group was twice as large, which increases the ability to detect effects in the Dep+Anx versus the Dep group. Fourth, for dimensional measures, the interpretation of the affective modulation of startle is limited as we see similar effects across valence, but we suggest that when the groups are collapsed the blunting effect of depression on overall startle overshadows affective modulation. However, we strongly feel that propensity matching within sub-types of MDD aims to address a crucial gap in the literature and can contribute to solving the crisis of heterogeneity in MDD research. Blunted startle could be related to a single impairment downstream from these systems in non-anxious depression. For example, a sensory deficit (Serafini et al., 2017) or a motor deficit (Sobin & Sackeim, 1997). Sensory deficits are difficult to investigate with the current data but our lack of valence independent group differences on RTs suggests that these groups may not have a single downstream motor deficit that could drive our startle findings. In addition, a recent review (Boecker & Pauli, 2019) suggests that the appetitive and defensive systems operate independently and are affected in opposing ways by anxiety and depression. Although behavioral inhibition was significantly different between the groups it was not associated with startle, although we would argue that this is unsurprising as behavioral inhibition is more related to cognitive arousal. The ASI was the best measure of anxious arousal that we had in our self-report data. Future studies of threat sensitivity should use more specific self-report scales than the ASI, for example the Trait Fear Scale (Kramer et al., 2020). Finally, there were no group differences on heart rate variability, although the emotional reactivity task was not designed to probe this.

In sum, these findings suggest that those with anxious and non-anxious MDD have distinct neurocognitive profiles and thus, may require different treatment approaches.

Exaggerated defensive responses in anxious MDD may promote avoidance behavior, whereas a lack of positive affect may be driving the same outcome in non-anxious MDD. The preservation of positive affective modulation suggests that anxious MDD is not as simple as *depression plus anxiety* and that there may be an interactive effect of depression and anxiety that could be key to understanding and treating this comorbidity. To our knowledge, this is the first time that findings on preservation of the appetitive system in anxious MDD have been presented; enabled by our propensity matching approach using both categorical and dimensional analyses. This appetitive system preservation may be a useful tool for developing strategies to regulate negative affect and hyperarousal in this large patient group.

## Data Availability

All data produced in the present study are available upon reasonable request to the authors

## Acknowledgements

This work has been supported in part by The William K. Warren Foundation and the National Institute of General Medical Sciences Center Grant Award Number 1P20GM121312. The content is solely the responsibility of the authors and does not necessarily represent the official views of the National Institutes of Health.

The ClinicalTrials.gov identifier for the clinical protocol associated with data published in the current paper is NCT02450240, “Latent Structure of Multi-level Assessments and Predictors of Outcomes in Psychiatric Disorders”.

The authors thank all the research participants and wish to acknowledge the contributions of Tulsa 1000 Investigators towards the collecting and organizing of data.

Maria Ironside, Rayus Kuplicki, Jennifer Stewart, Jonathan Savitz, Robin Aupperle, and Martin Paulus receive funding from the National Institute of General Medical Sciences (NIGMS) center grant P20GM121312; Maria Ironside has additional funding from National Institute of Mental Health (NIMH; R01MH132565). Sahib Khalsa has grant funding from the NIMH (K23MH112949, R01MH127225); Robin Aupperle has additional grant funding from NIMH (K23MH108707; R01MH123691); Jennifer Stewart has additional grant funding from the National Institute of Drug Abuse (NIDA) (R01DA050677), Rayus Kuplicki has additional funding from NIDA (R01DA050677); and Martin Paulus has additional grant funding from the National Institute of Drug Abuse (U01DA041089, R01DA050677).

## Author Contribution Statement

Maria Ironside: Conceptualization, formal analysis, writing – original draft

Rayus Kuplicki: Formal analysis, methodology, software, writing – review and editing

Ebony A. Walker: Data curation

Tate Poplin: Data curation, formal analysis

Cheldyn M. Ramsey: Data curation

Katherine L. Forthman: Formal analysis, software, writing – review and editing

Melissa E. Nestor: Software

Robin L. Aupperle: Conceptualization, writing – review and editing

Salvador M. Guinjoan: Writing – review and editing

Sahib S. Khalsa: Conceptualization, writing – review and editing

Jonathan Savitz: Conceptualization, writing – review and editing

Jennifer L. Stewart: Conceptualization, writing – review and editing

Teresa A. Victor: Conceptualization, writing – review and editing

Martin P. Paulus: Conceptualization, supervision, methodology, funding acquisition, writing – review and editing

## Notes

### Competing Interest Statement

Dr. Paulus is an advisor to Spring Care, Inc., a behavioral health startup, he has received royalties for an article about methamphetamine in UpToDate; Dr. Ironside reports no financial relationships with commercial interests; Dr. Kuplicki reports no financial relationships with commercial interests; Ms. Walker reports no financial relationships with commercial interests; Mr. Poplin reports no financial relationships with commercial interests; Ms. Ramsey reports no financial relationships with commercial interests; Ms. Forthman reports no financial relationships with commercial interests; Ms. Nestor reports no financial relationships with commercial interests; Dr. Aupperle reports no financial relationships with commercial interests; Dr. Guinjoan reports no financial relationships with commercial interests; Dr. Khalsa reports no financial relationships with commercial interests; Dr. Savitz reports no financial relationships with commercial interests; Dr. Stewart reports no financial relationships with commercial interests; Dr. Victor reports no financial relationships with commercial interests.

### Author Declarations

Ethical approval was obtained from Western Institutional Review Board T1000 protocol #20142082.

### Summary of Updates

This manuscript added justification of the comparison between two clinical groups because of healthy comparisons lying in the middle of the two clinical groups in terms of hyperarousal. HRV analysis was also added to this revision and the associated co-author added.

## References

1. Bar-Haim, Y., Lamy, D., Pergamin, L., Bakermans-Kranenburg, M. J., & van, I. M. H. (2007). Threat-related attentional bias in anxious and nonanxious individuals: a meta-analytic study. Psychol Bull, 133(1), 1–24. 10.1037/0033-2909.133.1.1

2. Blumenthal, T. D., Cuthbert, B. N., Filion, D. L., Hackley, S., Lipp, O. V., & van Boxtel, A. (2005). Committee report: Guidelines for human startle eyeblink electromyographic studies. Psychophysiology, 42(1), 1–15. 10.1111/j.1469-8986.2005.00271.x

3. Boecker, L., & Pauli, P. (2019). Affective startle modulation and psychopathology: Implications for appetitive and defensive brain systems. Neurosci Biobehav Rev, 103, 230–266. 10.1016/j.neubiorev.2019.05.019

4. Bradley, M. M., Cuthbert, B. N., & Lang, P. J. (1990). Startle reflex modification: emotion or attention? Psychophysiology, 27(5), 513–522. 10.1111/j.1469-8986.1990.tb01966.x

5. Buchanan, T. W., Tranel, D., & Adolphs, R. (2004). Anteromedial temporal lobe damage blocks startle modulation by fear and disgust. Behav Neurosci, 118(2), 429–437. 10.1037/0735-7044.118.2.429

6. Clark, L. A., & Watson, D. (1991). Tripartite model of anxiety and depression: psychometric evidence and taxonomic implications. J Abnorm Psychol, 100(3), 316–336. 10.1037//0021-843x.100.3.316

7. Cook, E. W., 3rd, Davis, T. L., Hawk, L. W., Spence, E. L., & Gautier, C. H. (1992). Fearfulness and startle potentiation during aversive visual stimuli. Psychophysiology, 29(6), 633–645. 10.1111/j.1469-8986.1992.tb02038.x

8. Cook, E. W., 3rd, Hawk, L. W., Jr., Davis, T. L., & Stevenson, V. E. (1991). Affective individual differences and startle reflex modulation. J Abnorm Psychol, 100(1), 5–13. 10.1037//0021-843x.100.1.5

9. Davidson, R. J. (1998). Affective style and affective disorders: Perspectives from affective neuroscience. Cognition & Emotion, 12(3), 307–330.

10. Davis, M., Walker, D. L., & Lee, Y. (1999). Neurophysiology and Neuropharmacology of Startle and Its Affective Modification. In A. H. Bohmelt, A. M. Schell, & M. E. Dawson (Eds.), Startle Modification (pp. 95–113). Cambridge University Press. 10.1017/cbo9780511665523.007

11. Etkin, A., & Schatzberg, A. F. (2011). Common abnormalities and disorder-specific compensation during implicit regulation of emotional processing in generalized anxiety and major depressive disorders. Am J Psychiatry, 168(9), 968–978. 10.1176/appi.ajp.2011.10091290

12. Fava, M., Rush, A. J., Alpert, J. E., Balasubramani, G. K., Wisniewski, S. R., Carmin, C. N., Biggs, M. M., Zisook, S., Leuchter, A., Howland, R., Warden, D., & Trivedi, M. H. (2008). Difference in treatment outcome in outpatients with anxious versus nonanxious depression: a STAR*D report. Am J Psychiatry, 165(3), 342–351. 10.1176/appi.ajp.2007.06111868

13. Forthman, K. L. (2019). optLog. In https://github.com/kforthman/optLog

14. Garner, M., Clarke, G., Graystone, H., & Baldwin, D. S. (2011). Defensive startle response to emotional social cues in social anxiety. Psychiatry Res, 186(1), 150–152. 10.1016/j.psychres.2010.07.055

15. Gaspersz, R., Lamers, F., Kent, J. M., Beekman, A. T. F., Smit, J. H., van Hemert, A. M., Schoevers, R. A., & Penninx, B. (2017). Anxious distress predicts subsequent treatment outcome and side effects in depressed patients starting antidepressant treatment. J Psychiatr Res, 84, 41–48. 10.1016/j.jpsychires.2016.09.018

16. Globisch, J., Hamm, A. O., Esteves, F., & Ohman, A. (1999). Fear appears fast: temporal course of startle reflex potentiation in animal fearful subjects. Psychophysiology, 36(1), 66–75. 10.1017/s0048577299970634

17. Grillon, C., & Ernst, M. (2020). A way forward for anxiolytic drug development: Testing candidate anxiolytics with anxiety-potentiated startle in healthy humans. Neurosci Biobehav Rev, 119, 348–354. 10.1016/j.neubiorev.2020.09.024

18. He, C., Gong, L., Yin, Y., Yuan, Y., Zhang, H., Lv, L., Zhang, X., Soares, J. C., Zhang, H., Xie, C., & Zhang, Z. (2019). Amygdala connectivity mediates the association between anxiety and depression in patients with major depressive disorder. Brain Imaging Behav, 13(4), 1146–1159. 10.1007/s11682-018-9923-z

19. Ionescu, D. F., Niciu, M. J., Richards, E. M., & Zarate, C. A., Jr. (2014). Pharmacologic treatment of dimensional anxious depression: a review. Prim Care Companion CNS Disord, 16(3). 10.4088/PCC.13r01621

20. Ironside, M., DeVille, D. C., Kuplicki, R. T., Burrows, K. P., Smith, R., Teed, A. R., Paulus, M. P., & Khalsa, S. S. (2023). The unique face of comorbid anxiety and depression: increased interoceptive fearfulness and reactivity. Frontiers in behavioral neuroscience.

21. Kaiser, R. H., Andrews-Hanna, J. R., Wager, T. D., & Pizzagalli, D. A. (2015). Large-Scale Network Dysfunction in Major Depressive Disorder: A Meta-analysis of Resting-State Functional Connectivity. JAMA Psychiatry, 72(6), 603–611. 10.1001/jamapsychiatry.2015.0071

22. Kaviani, H., Gray, J. A., Checkley, S. A., Raven, P. W., Wilson, G. D., & Kumari, V. (2004). Affective modulation of the startle response in depression: influence of the severity of depression, anhedonia, and anxiety. J Affect Disord, 83(1), 21–31. 10.1016/j.jad.2004.04.007

23. Kessler, R. C., Sampson, N. A., Berglund, P., Gruber, M. J., Al-Hamzawi, A., Andrade, L., Bunting, B., Demyttenaere, K., Florescu, S., de Girolamo, G., Gureje, O., He, Y., Hu, C., Huang, Y., Karam, E., Kovess-Masfety, V., Lee, S., Levinson, D., Medina Mora, M. E., … Wilcox, M. A. (2015). Anxious and non-anxious major depressive disorder in the World Health Organization World Mental Health Surveys. Epidemiol Psychiatr Sci, 24(3), 210–226. 10.1017/S2045796015000189

24. Kim, M. J., Loucks, R. A., Palmer, A. L., Brown, A. C., Solomon, K. M., Marchante, A. N., & Whalen, P. J. (2011). The structural and functional connectivity of the amygdala: from normal emotion to pathological anxiety. Behav Brain Res, 223(2), 403–410. 10.1016/j.bbr.2011.04.025

25. Koch, M., Schmid, A., & Schnitzler, H.-U. (1996). Pleasure-attenuation of startle is disrupted by lesions of the nucleus accumbens. NeuroReport, 7(8), 1442–1446. https://journals.lww.com/neuroreport/Fulltext/1996/05310/Pleasure_attenuation_of_startle_is_disrupted_by.24.aspx

26. Kramer, M. D., Patrick, C. J., Hettema, J. M., Moore, A. A., Sawyers, C. K., & Yancey, J. R. (2020). Quantifying Dispositional Fear as Threat Sensitivity: Development and Initial Validation of a Model-Based Scale Measure. Assessment, 27(3), 533–546. 10.1177/1073191119837613

27. Kuhn, M., Wendt, J., Sjouwerman, R., Buchel, C., Hamm, A., & Lonsdorf, T. B. (2020). The Neurofunctional Basis of Affective Startle Modulation in Humans: Evidence From Combined Facial Electromyography and Functional Magnetic Resonance Imaging. Biol Psychiatry, 87(6), 548–558. 10.1016/j.biopsych.2019.07.028

28. Lang, P. J. (1995). The emotion probe. Studies of motivation and attention. Am Psychol, 50(5), 372–385. 10.1037//0003-066x.50.5.372

29. Lang, P. J., Bradley, M. M., & Cuthbert, B. N. (1990). Emotion, attention, and the startle reflex. Psychol Rev, 97(3), 377–395. https://www.ncbi.nlm.nih.gov/pubmed/2200076

30. Lang, P. J., Bradley, M. M., & Cuthbert, B. N. (1999). International affective picture system (IAPS): Instruction manual and affective ratings. The center for research in psychophysiology, University of Florida.

31. MATLAB. (Version 7.10.0 (R2019a)) The MathWorks Inc.

32. McTeague, L. M., & Lang, P. J. (2012). The anxiety spectrum and the reflex physiology of defense: from circumscribed fear to broad distress. Depress Anxiety, 29(4), 264–281. 10.1002/da.21891

33. McTeague, L. M., Rosenberg, B. M., Lopez, J. W., Carreon, D. M., Huemer, J., Jiang, Y., Chick, C. F., Eickhoff, S. B., & Etkin, A. (2020). Identification of Common Neural Circuit Disruptions in Emotional Processing Across Psychiatric Disorders. Am J Psychiatry, 177(5), 411–421. 10.1176/appi.ajp.2019.18111271

34. Mobbs, D., Marchant, J. L., Hassabis, D., Seymour, B., Tan, G., Gray, M., Petrovic, P., Dolan, R. J., & Frith, C. D. (2009). From threat to fear: the neural organization of defensive fear systems in humans. J Neurosci, 29(39), 12236–12243. 10.1523/JNEUROSCI.2378-09.2009

35. Mobbs, D., Petrovic, P., Marchant, J. L., Hassabis, D., Weiskopf, N., Seymour, B., Dolan, R. J., & Frith, C. D. (2007). When fear is near: threat imminence elicits prefrontal-periaqueductal gray shifts in humans. Science, 317(5841), 1079–1083. 10.1126/science.1144298

36. Penninx, B. W., Nolen, W. A., Lamers, F., Zitman, F. G., Smit, J. H., Spinhoven, P., Cuijpers, P., de Jong, P. J., van Marwijk, H. W., van der Meer, K., Verhaak, P., Laurant, M. G., de Graaf, R., Hoogendijk, W. J., van der Wee, N., Ormel, J., van Dyck, R., & Beekman, A. T. (2011). Two-year course of depressive and anxiety disorders: results from the Netherlands Study of Depression and Anxiety (NESDA). J Affect Disord, 133(1-2), 76–85. 10.1016/j.jad.2011.03.027

37. Powers, S. I., Laurent, H. K., Gunlicks-Stoessel, M., Balaban, S., & Bent, E. (2016). Depression and anxiety predict sex-specific cortisol responses to interpersonal stress. Psychoneuroendocrinology, 69, 172–179. 10.1016/j.psyneuen.2016.04.007

38. R Core Team. (2020). R: A language and environment for statistical computing. . In R Foundation for Statistical Computing. https://www.R-project.org/.

39. Rottenberg, J., Gross, J. J., & Gotlib, I. H. (2005). Emotion context insensitivity in major depressive disorder. Journal of abnormal psychology, 114(4), 627.

40. Serafini, G., Gonda, X., Canepa, G., Pompili, M., Rihmer, Z., Amore, M., & Engel-Yeger, B. (2017). Extreme sensory processing patterns show a complex association with depression, and impulsivity, alexithymia, and hopelessness. Journal of Affective Disorders, 210, 249–257. 10.1016/j.jad.2016.12.019

41. Shaffer, F., & Ginsberg, J. P. (2017). An overview of heart rate variability metrics and norms. Frontiers in public health, 258.

42. Sheehan, D. V., Lecrubier, Y., Sheehan, K. H., Amorim, P., Janavs, J., Weiller, E., Hergueta, T., Baker, R., & Dunbar, G. C. (1998). The Mini-International Neuropsychiatric Interview (M.I.N.I.): the development and validation of a structured diagnostic psychiatric interview for DSM-IV and ICD-10. J Clin Psychiatry, 59 Suppl 20(20), 22–33;quiz 34-57. https://www.ncbi.nlm.nih.gov/pubmed/9881538

43. Sobin, C., & Sackeim, H. A. (1997). Psychomotor symptoms of depression. American Journal of Psychiatry, 154(1), 4–17.

44. Temple, R. O., & Cook, E. W. (2007). Anxiety and defensiveness: Individual differences in affective startle modulation. Motivation and Emotion, 31(2), 115–123.

45. Vaidyanathan, U., Patrick, C. J., & Bernat, E. M. (2009). Startle reflex potentiation during aversive picture viewing as an indicator of trait fear. Psychophysiology, 46(1), 75–85. 10.1111/j.1469-8986.2008.00751.x

46. Victor, T. A., Khalsa, S. S., Simmons, W. K., Feinstein, J. S., Savitz, J., Aupperle, R. L., Yeh, H. W., Bodurka, J., & Paulus, M. P. (2018). Tulsa 1000: a naturalistic study protocol for multilevel assessment and outcome prediction in a large psychiatric sample. BMJ open, 8(1), e016620. 10.1136/bmjopen-2017-016620

47. Vollmer, M. (2019). HRVTool–an open-source matlab toolbox for analyzing heart rate variability. 2019 Computing in Cardiology (CinC),

48. Williams, L. M. (2017). Defining biotypes for depression and anxiety based on large-scale circuit dysfunction: a theoretical review of the evidence and future directions for clinical translation. Depress Anxiety, 34(1), 9–24. 10.1002/da.22556

49. Zhao, K., Liu, H., Yan, R., Hua, L., Chen, Y., Shi, J., Lu, Q., & Yao, Z. (2017). Cortical thickness and subcortical structure volume abnormalities in patients with major depression with and without anxious symptoms. Brain and behavior, 7(8), e00754. 10.1002/brb3.754

50. Zhao, Q.-Y., Luo, J.-C., Su, Y., Zhang, Y.-J., Tu, G.-W., & Luo, Z. (2021). Propensity score matching with R: conventional methods and new features. Annals of translational medicine, 9(9), 812–812. 10.21037/atm-20-3998

