## supplemental information for "The unique face of anxious depression: Exaggerated threat but preserved positive valence reactivity"

**Supplemental Methods:**

First 500 of T1000 sample

N = 500

EMG data collected

N = 478

EMG QC passed

N = 418

MINI: Non-anxious depression

N = 62

MINI: Anxious depression

N = 149

Propensity matched

Anxious depression

N = 124

Propensity matched

Non-anxious depression

N = 62

Excluded (N = 207)

Other diagnoses:

*Anxiety N = 16*

*Substance use N = 125*

*Eating disorder N = 15*

Missing self-report data:

*Non-anxious depression N = 1*

Excluded (N = 60 )

Poor quality EMG data

Healthy comparisons

N = 50

**Supplemental Figure S1: CONSORT diagram**

EMG: Electromyography; QC: Quality Control; MINI: MINI International Neuropsychiatric interview

**Parent Study Methods:**

The following supplemental materials summarizes the methods from the parent study. Full details are in the protocol paper (Victor et al., 2018).

**Parent Study Participants:** We collected datasets on a total of 1050 participants with approximately 506 mood and/or anxiety, 330 substance use, 54 eating disorder and 160 mentally and physically healthy control participants. In order to obtain at least 1000 participants who completed the year-long study, we enrolled 1272 participants between January 2015 and December 2018. Participants were between 18 and 55 years of age and had a body mass index between 17-38kg/m^2^. Participants were either referred from local treatment facilities or seeking treatment for anxiety and/or depressive symptoms, problems related to substance use, or problems related to eating behavior. As part of the inclusion criteria, mood/anxiety, substance, and eating disorder participants must have also screened positive for these conditions as indicated by a score on the Patient Health Questionnaire (PHQ-9) ≥ 10 and/or Overall Anxiety Severity and Impairment Scale (OASIS) ≥ 8, (DAST-10) score > 2 or Sick, Control, One, Fat, Food Questionnaire eating disorder screen (SCOFF) score ≥ 2. Participants who met criteria for one primary domain could also screen positive for one of the other study domains. Healthy control participants screened negative for these inclusion measures.

**Parent Study Design:**
The study’s dependent variables focus on the *positive and negative valence systems, cognition, and arousal/interoception domains* proposed by the RDoC (Health, 2011a, 2011b). Using self-report, behavior, physiology, neural circuit, cell, molecule, and gene unit of analysis measures, these constructs were applied to a clinical population of individuals with dysregulation of affect, substance use, and eating behavior recruited from treatment providers across different sites in the community. Participants underwent a multi-level assessment based on the RDoC approach that consists of (a) a standardized diagnostic assessment, (b) self-report questionnaires assessing the positive and negative valence domains as well as interoception, (c) behavioral tasks assessing positive and negative valence, cognition, and interoception, (d) physiological measurements consisting of skin conductance, facial emotion expression monitoring, heart rate, respiration and eye-blink startle response, (e) functional magnetic resonance imaging focusing on reward-related processing, fear conditioning and extinction, cognitive control and inhibition, and interoceptive processing, (f) biomarker assessment, (g) microbiome assessment, (h) blood to derive induced pluripotent stem cells (IPS), (i) and genetic as well as epigenetic assessments. Subsequently, these individuals were followed up quarterly and for one year. At months 3, 6, and 9, only self-report assessments were collected, and the participants were re-assessed using a multi-domain assessment of functioning, which included: (a) symptom severity and duration, (b) subjective well-being, (c) psychosocial function, (c) occupational function, (d) physical health, (e) utilization of mental health resources (treatment), and (f) adherence to treatment.

The workflow schematic in Figure 1 describes the overall outline of the T-1000 study and the measures obtained at different points in time.

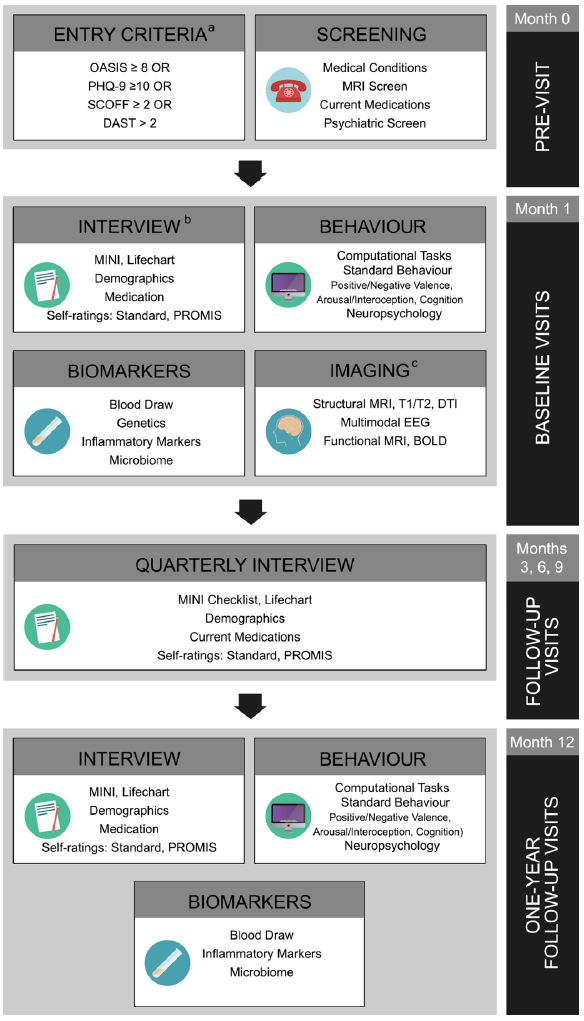

**Supplemental Figure S2: Tulsa 1000 workflow schematic.** BOLD, blood oxygen level-dependent; DAST, drug abuse screening test; DTI, diffusion tensor imaging; EEG, electroencephalogram; MINI, Mini International Neuropsychiatric Interview; OASIS, Overall Anxiety Severity and Impairment Scale; PHQ-9, Patient Health Questionnaire; PROMIS, Patient-Reported Outcome Measurement Information System; SCOFF, Sick,Control, One, Fat, Food Questionnaire; T1/T2, T1-weighted (longitudinal relaxation time) and T2-weighted (transverse relaxation time). Reproduced with permission from (Victor et al., 2018).

**Supplemental Results:**

***Image ratings:***

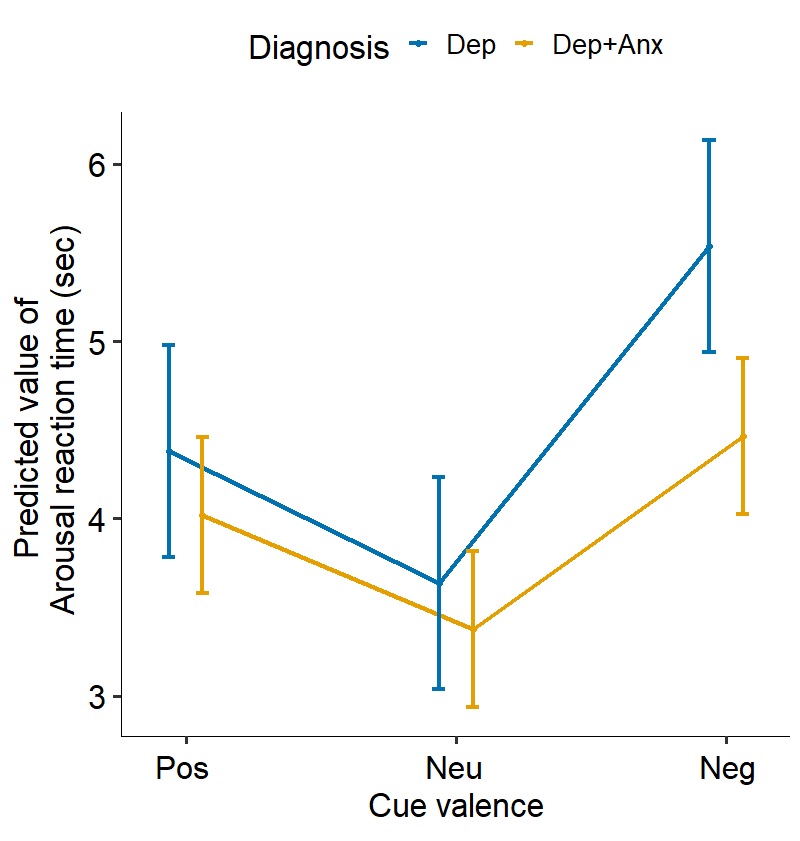

**Supplemental Figure S3:** Increased reaction time for arousal ratings for negative images depression versus comorbid depression and anxiety. Error bars represent the 95% confidence interval of the model predicted value.

***Healthy control comparison:*** Although the hypotheses of the study are somewhat obscured by a healthy comparisons as they are predicted to lie between the two clinical groups, for completeness, follow-up analyses compared propensity matched Dep and Dep+Anx groups to healthy comparisons (HC; N = 50). For the Dep+Anx vs HC analyses there was no significant main effect of group or any valence X group interaction (all *p* > 0.8). For the Dep vs HC analyses mixed effects linear regression results showed a trend *valence* X *group* interaction (F(1,100) 3.105, *p* = 0.08, R2 = 0.030). Planned contrasts showed that the Dep group had no modulation of startle response from valence (pairwise comparisons; all *p* > 0.6), whereas the HC group showed negative potentiation compared to positive (negative > positive) (t(100) = 2.724, p = 0.008) and positive attenuation compared to neutral (positive < neutral) (t(294) = -2.148, p = 0.03). There were no differences between the groups on appetitive, neutral or aversive cues (pairwise comparisons; all *p* > 0.1). A post-hoc analysis, comparing all groups in one mixed-model, the *valence* X *group* interaction was reduced to a trend (*p* = 0.07). However, as outlined in the introduction, comparisons on measures of arousal between these two groups and healthy comparisons may not be helpful as they would be expected to lie in between the hyperaroused Dep+Anx group and the blunted Dep group.

**Supplemental References:**

Health, N. I. o. M. (2011a). Negative Valence Systems: Workshop Proceedings. Retrieved from <http://www.nimh.nih.gov/research-funding/rdoc/negative-valence-systems-workshop-proceedings.shtml>

Health, N. I. o. M. (2011b). Positive Valence Systems: Workshop Proceedings. Retrieved from <http://www.nimh.nih.gov/research-funding/rdoc/positive-valence-systems-workshop-proceedings.shtml>

Victor, T. A., Khalsa, S. S., Simmons, W. K., Feinstein, J. S., Savitz, J., Aupperle, R. L., . . . Paulus, M. P. (2018). Tulsa 1000: a naturalistic study protocol for multilevel assessment and outcome prediction in a large psychiatric sample. *BMJ open, 8*(1), e016620. doi:10.1136/bmjopen-2017-016620
